# Relative effectiveness of booster vs. 2-dose mRNA Covid-19 vaccination in the Veterans Health Administration: Self-controlled risk interval analysis

**DOI:** 10.1101/2022.03.17.22272555

**Authors:** Caroline Korves, Hector S. Izurieta, Jeremy Smith, Gabrielle M. Zwain, Ethan I. Powell, Abirami Balajee, Kathryn Ryder, Yinong Young-Xu

## Abstract

**Importance:** Previous studies have analyzed effectiveness of booster mRNA Covid-19 vaccination and compared it with 2-dose primary series for both Delta and Omicron variants. Observational studies that estimate effectiveness by comparing outcomes among vaccinated and unvaccinated individuals may suffer from residual confounding and exposure misclassification.

**Objective:** To estimate relative effectiveness of booster vaccination versus the 2-dose primary series with self-controlled study design

**Design, Setting and Participants:** We used the Veterans Health Administration (VHA) Corporate Data Warehouse to identify U.S. Veterans enrolled in care ≥2 years who received the 2-dose primary mRNA Covid-19 vaccine series and a mRNA Covid-19 booster following expanded recommendation for booster vaccination, and who had a positive SARS-CoV-2 test during the Delta (9/23/2021-11/30/2021) or Omicron (1/1/22-3/1/22) predominant period. Among them, we conducted a self-controlled risk interval (SCRI) analysis to compare odds of SARS-CoV-2 infection during a booster exposure interval versus a control interval.

**Exposures:** control interval (days 4-6 post-booster vaccination, presumably prior to gain of booster immunity), and booster exposure interval (days 14-16 post-booster vaccination, presumably following gain of booster immunity)

**Outcomes and Measures:** Positive PCR or antigen SARS-CoV-2 test. Separately for Delta and Omicron periods, we used conditional logistic regression to calculate odds ratios (OR) of a positive test for the booster versus control interval and calculated relative effectiveness of booster versus 2-dose primary series as (1-OR)*100. The SCRI approach implicitly controlled for time-fixed confounders.

**Results:** We found 42 individuals with a positive SARS-CoV-2 test in the control interval and 14 in the booster exposure interval during Delta period, and 137 and 66, respectively, in Omicron period. For the booster versus 2-dose primary series, the odds of infection were 70% (95%CI: 42%, 84%) lower during the Delta period and 56% (95%CI: 38%, 67%) lower during Omicron. Results were similar for ages <65 and ≥65 years in the Omicron period. In sensitivity analyses among those with prior Covid-19 history, and age stratification, ORs were similar to the main analysis.

**Conclusions:** Booster vaccination was more effective relative to a 2-dose primary series, the relative effectiveness was consistent across age groups and was higher during the Delta predominant period than during the Omicron period.

## INTRODUCTION

Following the Food and Drug Administration (FDA) Emergency Use Authorization (EUA) for the mRNA Covid-19 vaccines, several studies demonstrated high vaccine effectiveness (VE) for these vaccines in real-world settings in the United States (US).^1-5^ By July 2021, Delta became the predominant circulating SARS-CoV-2 variant in the US, and reports of breakthrough infections rose along with questions regarding waning immunity of the mRNA vaccines.^6, 7^ These occurrences prompted the Advisory Committee on Immunization Practices to recommend booster vaccination and, starting in September 2021, to allow a booster dose for a larger portion of the population.^8-10^

Booster vaccine effectiveness (VE) for mRNA vaccines from real world settings in and outside the US have shown lower VE against infection with Omicron than Delta variants. A study in southern California for December 2021 reported lower booster vaccine effectiveness (VE) against infection for Omicron (62.5%) than Delta (95.2%).^11^ A CDC-led study across the US reported booster VE against infection declined from 93% to 80% from the period of Delta predominance to Omicron emergence^12^, and a separate CDC-led study found similar declines of 94% to 82% among patients tested in emergency department/urgent care encounters.^13^ In Israel the recommendation for booster vaccination preceded the US;^14^ analyses of patient data from August-September 2021, when Delta was predominant, showed risk of infection was 12 times lower among those boosted versus those not boosted^14^.^14, 15^

Since observational studies contribute to our understanding of VE, and policy decisions and scientific recommendations are based in part upon observational studies, it is important to have confidence in the findings. Observational studies from different settings and pandemic time periods contribute to our understanding of VE. All observational studies must account for confounding, and studies of VE must account for differences between vaccinated and unvaccinated people that may contribute to different risk of infection. Observational studies must also minimize misclassification of vaccination status which can bias results. To reduce such confounding, we utilized a self-controlled study design that implicitly accounts for time-fixed confounders. Using data from veterans who were recorded as fully vaccinated with two doses of mRNA Covid-19 vaccines and later boosted, we quantified odds of infection for booster vaccination versus the 2-dose primary series. We used data from the Veterans Health Administration (VHA) population, which offers a unique opportunity to better our understanding of the booster vaccination across the US. Because of the ability to analyze data in near real-time, it was possible to assess the effectiveness of the booster with the establishment of Omicron as the predominant variant.

## METHODS

### Data Sources

The VHA is the largest integrated health care system in the U.S., providing comprehensive care to over nine million veterans at more than 171 medical centers and 1,112 outpatient sites of care.^16^ Electronic medical record data from the VHA Corporate Data Warehouse (CDW) were analyzed. We used publicly available data from the Centers for Disease Control and Prevention on weekly monitoring of variant proportions in the US to identify the predominant SARS-CoV-2 variants for time periods in the study.^17^

### Study Design

We used a self-controlled risk interval (SCRI) study design, a variation of the self-controlled case series (SCCS) design.^18^ The method can be used for non-recurrent events when the risk of occurrence over the study period is 10% or less,^19^ which was the case with the current study. With this design, only cases are included in the analysis, and periods of exposure and non-exposure around an event of interest are identified. The SCRI design is beneficial when studying an exposure where identifying an unbiased comparable cohort is difficult, as in our case in which people with booster vaccination are potentially different from the non-boosted in multiple measurable and unmeasurable ways, thus increasing the risk of residual confounding for cohort or case-control designs despite adjustment. In SCRI, time-fixed confounders are implicitly adjusted for because the risk and control intervals belong to the same individual and, accordingly, the analysis is matched. We shortened the length of intervals for analysis to be segments of the exposure and non-exposure periods to decrease time-varying confounding as well. Furthermore, by restricting the analysis only to patients with recorded booster vaccination we avoided misclassification of booster vaccination status.

A prior study by Bar-On *et al*^*15*^ of the BNT162b2 messenger mRNA vaccine, assumed that (anamnestic) immune response would start at around day seven post-booster vaccination and that testing is likely to follow infection by 5 days (incubation period). They^15^ selected days >12 post-booster vaccination as the time period during which the vaccinee should have already benefited from the booster dose. Following a similar logic, we used 4-6 days post-vaccination as the “control interval” to represent non-boosted exposure status, assuming a positive SARS-CoV-2 test during this time would likely reflect infection that occurred prior to the effect of booster vaccination. Also *a priori* we selected a “booster exposure” interval of days 14-16 post-booster vaccination, of the same length (3 days) as the control interval, specifically to represent a short interval within the presumed boosted effect time period that was also close (within a two-week period) of the control interval, minimizing the potential for large differences in community transmission of SARS-CoV-2 between the booster exposure and control intervals (Figure 1). The control and booster exposure intervals from the same individual formed a matched pair for analysis.

**Figure 1.**
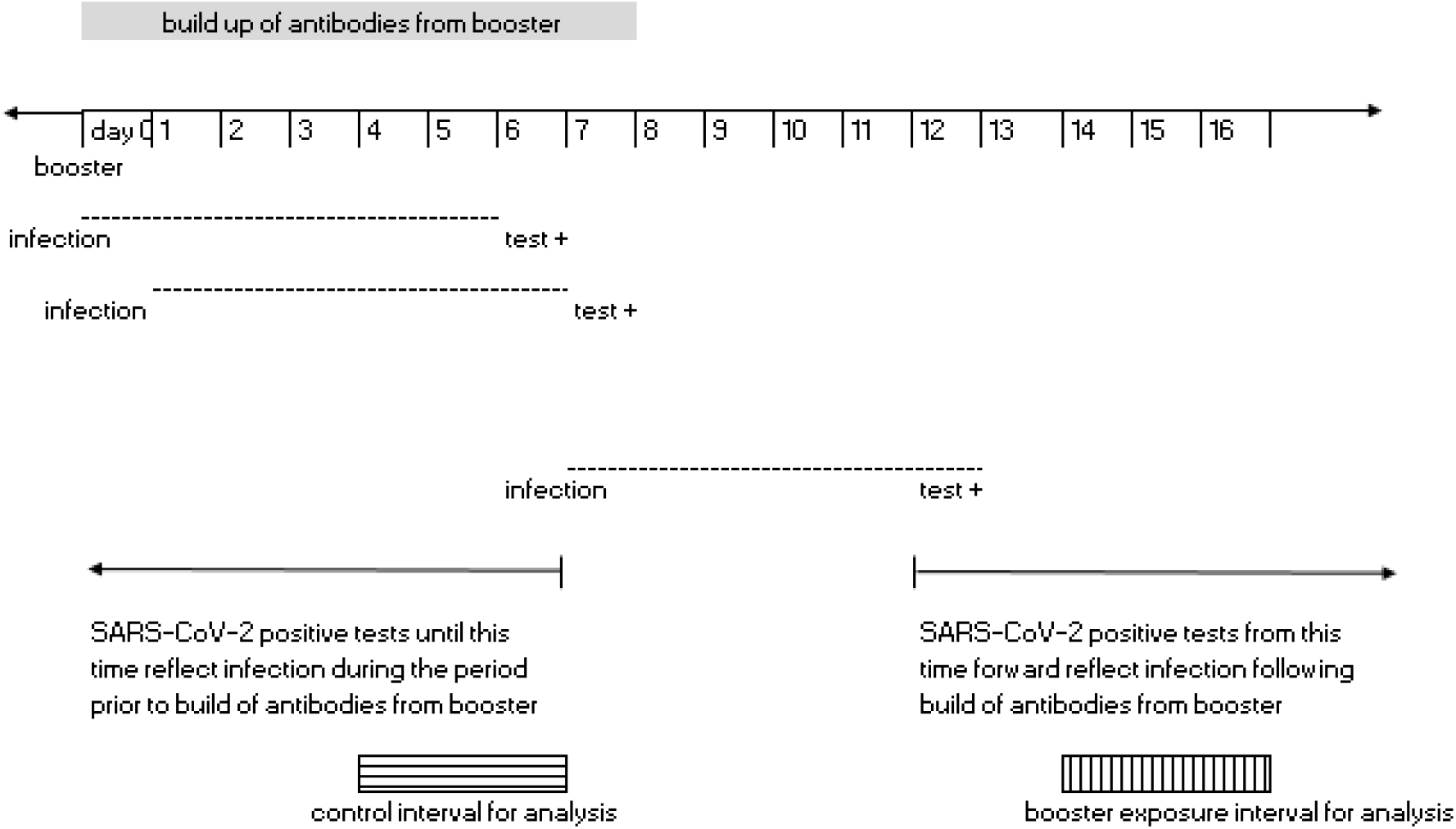
Booster exposure and control intervals for analysis

### Study Population

This analysis focused on VHA-enrolled veterans who were vaccinated and later boosted with an mRNA Covid-19 vaccine. The study population was restricted to veterans who received two Pfizer-BioNTech Covid-19 vaccines or two Moderna Covid-19 vaccines during December 14, 2020-August 1, 2021 and subsequently received a third mRNA vaccine (i.e., booster) of either mRNA vaccine type September 23, 2021 or thereafter. We restricted the population to vaccinees who received their 2-dose primary series by August 1, 2021 because we were interested in examining the booster effect during Delta and Omicron predominant periods through March 1 2022, and most individuals who received a 2-dose primary series after August 1, 2021 would not be recommended for booster vaccination given the recency of their 2-dose primary series. Vaccinees were veterans living in the US, enrolled in VHA ≥2 years prior to the vaccination era (December 14, 2020), and had ≥1 visit to a VHA facility in the prior 2 years. In the main analysis, we did not include those individuals who had a prior Covid-19 diagnosis, had a positive antigen test or a positive PCR test prior to booster vaccination. As a condition of the SCRI study design, only vaccine recipients who had a positive PCR or antigen SARS-CoV-2 PCR test during the control interval (i.e., days 4-6 post booster vaccination) or booster exposure interval (i.e., days 14-16 post booster vaccination) were included in the analysis. Tests from veterans who were hospitalized for more than one day at time of testing were excluded.

We classified SARS-CoV-2 tests during September 23, 2021-November 30, 2021 as being from the Delta-predominant period and those in January 1-March 1, 2022 from the Omicron-predominant period, based on the CDC’s data which showed nearly 100% of sequenced samples were Delta during August 2021-November 2021, and 90-100% were Omicron January 1-March 1, 2022.^17^ We excluded tests taken from veterans during December 2021 as the predominant variant shifted during this time. Tests had to be from individuals whose booster exposure interval and control interval fell within the variant-predominant period of analysis.

### Exposure, Outcome and Covariate Assessment

Using the positive SARS-CoV-2 test data from the population of vaccinees who met the study criteria, we classified individuals who had a positive test in the booster exposure or control intervals as cases and determined the exposure status of the case by whether the test date occurred in the booster exposure or control interval.

The SCRI design adjusts for confounding by matching an individual to themself close in time, minimizing or eliminating the need for additional adjustment. We did adjust for the booster exposure interval or control interval including a weekend day, as an individual’s likelihood of being tested could be different during a weekend. We included other variables in a descriptive analysis and used them to conduct stratified analyses.

We conducted a falsification analysis using the exposure of influenza vaccination (not on the same day as Covid-19 vaccination) at a VHA facility and compared risk of a positive SARS-CoV-2 test days 14-16 versus days 4-6 post influenza vaccination as we would expect that the risk of SARS-CoV-2 infection to be no different for these time periods around influenza vaccination.

### Statistical analysis

The population of vaccinees who met the study criteria for the Delta and Omicron periods were identified and described. Mean (standard deviation) and median (interquartile range) were reported for continuous variables, and frequency and proportions reported for categorical variables. In this SCRI analysis, we used conditional logistic regression to calculate the odds ratio (OR) (95% confidence interval [CI]) of a positive test for the booster exposure versus the control interval, and conducted analyses separately for the Delta predominant and Omicron predominant periods and for subgroups stratified by age (<65 years and ≥65 years). We estimated the relative effectiveness of the booster versus the 2-dose primary series as the percentage reduction in the odds of testing positive for the low-risk booster exposure interval versus the control interval ([1-OR]*100%).

### Sensitivity Analyses

Additional analyses were conducted including patients with a history of Covid-19 prior to booster vaccination so long as the last SARS-CoV-2 positive test was ≥90 days prior to the SARS-CoV-2 test following booster vaccination.

All analyses were conducted using SAS, version 9.4.

Approval: The study protocol was approved by the institutional review board of the VA Medical Center in White River Junction, VT and was granted an exemption of consent.

## RESULTS

A total of2,650,278 Veterans had completed the 2-dose primary series by August 1, 2021 and had not been boosted by September 23, 2021. Among them, 1,240,999 (47%) received a booster September 23, 2021-March 1, 2022. The median time from 2nd dose until booster was 247 days (range 59-415 IQR 227-270). Among those boosted in this time interval, 9,354 (0.75%) had a positive SARS-CoV-2 test after the booster, with 301 during the Delta period, 7,621 during the Omicron period, and the remaining in December 2021.

We restricted the analysis population to individuals with a SARS-CoV-2 positive test during the control or booster exposure intervals which included 56 individuals during the Delta period and 203 during the Omicron period. In the Delta period, there were 42 cases in the control interval, and 14 in the booster exposure interval. In the Omicron period, there were 137 cases in the control interval, and 66 in the booster exposure interval. The characteristics of the study population are shown in Table 1.

**Table 1.** Characteristics of individuals with positive SARS-CoV-2 test during Delta and Omicron periods

|  | Delta period (n=56) | Omicron period (n=203) |
| --- | --- | --- |
| Age, years, mean (median) | 71 (72) | 60 (61) |
| Sex, n(%) | * |  |
| Male |  | 178 (88) |
| Female |  | 25 (12) |
| Race/ethnicity, n(%) | * |  |
| Non-Hispanic white |  | 116 (57) |
| Non-Hispanic black |  | 67 (33) |
| Other/ unknown |  | 20 (10) |
| Rurality, n(%) |  |  |
| Rural | 21 (38) | 32 (21) |
| Urban | 35 (62) | 143 (73) |
| Booster manufacturer, n(%) |  |  |
| Pfizer | 31 (55) | 78 (38) |
| Moderna | 25 (45) | 125 (62) |
| 2-dose primary series manufacturer, n(%) |  |  |
| Pfizer | 31 (55) | 77 (38) |
| Moderna | 25 (45) | 126 (62) |
| *Reporting of number suppressed due to low value because of privacy concerns |  |  |

We presented the estimated relative effectiveness of the booster versus the 2-dose primary series in Table 2. There was a 70% (95%CI: 42%, 84%) reduction in the odds of testing positive in the booster exposure versus control interval during the Delta period. There was a reduction in the odds of testing positive for the booster exposure versus control interval during the Omicron period as well, but not as pronounced (56% (95%CI: 39%, 67%)). The estimated relative effectiveness of the booster versus 2-dose primary series was similar for individuals <65 years and ≥65 years during the Omicron period. We did not conduct stratified analysis for the Delta period given small numbers.

**Table 2.** Relative effectiveness of booster vaccination versus 2-dose primary series

|  | Number of positive SARS-CoV-2 tests |  |  |
| --- | --- | --- | --- |
|  | Booster exposure interval (days 14-16 post booster vaccination) | Control interval (days 4-6 post booster vaccination) | Relative effectiveness (95% CI) [reference: control interval]<sup>1</sup> |
| Delta period | 14 | 42 | 70% (42%, 84%) |
| Omicron period | 66 | 137 | 56% (39%, 67%) |
| <65 years old | 37 | 81 | 60% (34%, 74%) |
| ≥65 years old | 29 | 56 | 50% (20%, 68%) |
| Delta period: September 23, 2021-November 30, 2021; Omicron period: January 1, 2022- March 1, 2022. |  |  |  |
| <sup>1</sup>Relative effectiveness is the percentage reduction in the odds of testing positive for the booster exposure versus the control interval = $([1 - \text{OR}_{\text{odds of SARS-CoV-2 in booster exposure interval vs odds of SARS-CoV-2 in control interval}}] * 100\%)$. | | | |

In the falsification analysis we found no statistically significant association between the time interval around influenza vaccination and SARS-CoV-2 infection (OR of SARS-CoV-2 infection for days 14-16 vs days 4-6 post influenza vaccination: [OR=1.38 (95%: 0.89, 2.2)]).

For the sensitivity analysis, including veterans with a prior history of Covid-19, 59 had positive tests (cases) during the Delta period (43 in the control interval, 16 in the booster exposure interval) and 217 during the Omicron period (147 cases in the control interval, 70 cases in the booster exposure interval). The estimated reduction in odds of infection for booster exposure versus control interval was similar to the main analysis (Delta: 68% (95%CI: 39%, 83%); Omicron: 56% (95%CI: 41%, 68%)).

## DISCUSSION

In this study, booster vaccination was associated with a 70% reduction in infection compared with the 2-dose primary series during the Delta period and 56% reduction during Omicron. These differences were similar for those ages <65 and ≥65 years. Findings from this study align with prior observational studies showing that booster vaccination is associated with lower odds of infection compared with the 2-dose primary series^13^.^11-15, 20-22^ Prior studies ^11-13, 21^ showed greater estimated protection of booster vaccination versus the 2-dose primary series during the Delta compared with the Omicron period; while our findings were consistent with this, the difference in protection between the two periods was not as pronounced.

While most other studies evaluating booster vaccination relied on comparison with unvaccinated individuals, a limited number of studies^15, 20, 22^ have compared risk of infection for booster vaccination versus the 2-dose primary series (although not with a SCRI approach). One study conducted among individuals boosted in Israel during the Delta predominant period reported risk of infection was five times lower ≥12 days after the booster versus days 4-6 post-booster.^15^ A case-control study conducted in Israel during the Delta wave compared odds of infection for individuals with booster versus the 2-dose primary series, estimating an 83-87% reduction in risk for booster vaccination;^22^ this is comparable to the 70% relative effectiveness (OR=0.30) that we report for booster versus 2-dose primary for the Delta period. In a study of public health testing sites in the US, Accorsi *et al*^*21*^ estimated relative effectiveness was 84% for 3 doses versus 2 doses for confirmed Delta infection and 66% for Omicron infection, comparable to the 56% we report for the Omicron period.

The main strengths of this study are that by using a SCRI design we implicitly adjusted for all time-fixed confounders, and the very short interval we chose between windows minimizes the likelihood of all time varying confounding, including by differences in virus circulation. Also, our near real-time access to medical records allowed analysis of the booster dose for the Omicron period. Misclassification of vaccination status in other studies is often a concern as individuals may be vaccinated and boosted outside of their regular places of care and vaccination records may not be updated to reflect that for some time. We eliminated that risk by designing the study to only include those with known vaccination. While everyone in this study had three vaccinations at the VA, it is possible that some may have obtained additional vaccinations elsewhere. However, our population was limited to Veterans who routinely sought care at VHA facilities and given they had three vaccinations at a VHA facility they were unlikely to have also been vaccinated elsewhere.

We included days 4-6 post-vaccination as the control window, assuming immune response to vaccination will not occur so early (moreover, testing date is often subsequent to symptoms onset day); if some immune response to the booster dose was present during this control period, the bias would have been towards the null. Thus, our estimates of relative effectiveness of booster versus the 2-dose primary series may even be conservative. While the SCRI study design adjusts well for person-level factors that remain constant across exposure intervals, it is possible that individuals’ behaviors changed around time of vaccination which could alter infection risk. If individuals were more likely to reduce masking and other distancing measures following booster vaccination this could have underestimated the effect of booster vaccination during the booster exposure period when compared to the control period. Because the Delta and Omicron analysis periods occurred several months after the recommendation for third-dose booster vaccination for individuals with immunocompromising conditions, we assume that our analysis is most relevant to individuals without immunocompromising conditions as those with the conditions would have been boosted earlier.

Because we did not differentiate between symptomatic and asymptomatic disease, we did not measure effectiveness specifically for prevention of symptomatic COVID-19 disease. Although our study included tests from individuals regardless of symptoms, a majority of those tested for whom information was available had symptoms. Also, we did not consider the time interval between 2nd dose and booster dose; thus, we did not account for waning of effectiveness of the second dose. Therefore, our results show the relative effectiveness between a booster dose and 2 doses at the actual time at which the veterans received their booster dose (among veterans in this study, 247 days on average between 2nd dose and booster vaccination). Waning of effectiveness has been studied elsewhere, and one study reported VE against infection during the Delta period was 86% versus 76% for the 2-dose series when the time since second dose was <180 versus ≥180 days; the decrease in VE by time since vaccination during Omicron was similar (52% to 38%).^13^ Booster vaccination among individuals with such a long time since their 2nd dose likely had greater benefit from booster vaccination relative to those boosted closer in time to their 2nd dose. Also, our study only assessed the booster effect shortly after vaccination and, thus, did not address potential waning of protective immunity following the booster vaccination. Viral sequencing data were not available for each SARS-CoV-2 lab test included in this study, so we relied on CDC variant tracking data to classify cases as occurring during the Delta and Omicron predominant periods. To increase confidence in regard to the variant, we excluded the month of December 2021, during which the Delta-Omicron dominance was changing. Given that CDC data indicated near 100% Delta and Omicron predominance in our designated variant predominant periods, this was a reasonable assumption.

## CONCLUSION

This observational study indicates mRNA vaccine boosters were associated with a significant reduction in odds of infection relative to the 2-dose primary series during Omicron predominance, and more so during the Delta predominant period.

## Data Availability

Because data contain potentially identifying or sensitive patient information, all relevant data must be requested through the Department of Veterans Affairs at:
Research and Development Committee
VA Medical Center
163 Veterans Drive
White River Junction, VT 05009-0001

## Acknowledgements of research support for the study

This project was funded by the United States Food and Drug Administration through an interagency agreement with the Veterans Health Administration. Funding was also provided by the U.S. Department of Veterans Affairs (VA) Office of Rural Health.

